# Development and Prospective Validation of a Transparent Deep Learning Algorithm for Predicting Need for Mechanical Ventilation

**DOI:** 10.1101/2020.05.30.20118109

**Authors:** Supreeth P. Shashikumar, Gabriel Wardi, Paulina Paul, Paulina Paul, Morgan Carlile, Laura N Brenner, Kathryn A Hibbert, Crystal M. North, Shibani S. Mukerji, Gregory K. Robbins, Yu-Ping Shao, Atul Malhotra, M. Brandon Westover, Shamim Nemati

**Affiliations:** Department of Biomedical Informatics, University of California, San Diego; Department of Emergency Medicine, University of California, San Diego; Division of Pulmonary, Critical Care, and Sleep Medicine, University of California, San Diego; Division of Pulmonary and Critical Care Medicine, Massachusetts General Hospital, Boston; Department of Neurology, Massachusetts General Hospital, Boston; Division of Infectious Diseases, Massachusetts General Hospital, Boston

## Abstract

**IMPORTANCE:** Objective and early identification of hospitalized patients, and particularly those with novel coronavirus disease 2019 (COVID-19), who may require mechanical ventilation is of great importance and may aid in delivering timely treatment.

**OBJECTIVE:** To develop, externally validate and prospectively test a transparent deep learning algorithm for predicting 24 hours in advance the need for mechanical ventilation in hospitalized patients and those with COVID-19.

**DESIGN:** Observational cohort study

**SETTING:** Two academic medical centers from January 01, 2016 to December 31, 2019 (Retrospective cohorts) and February 10, 2020 to May 4, 2020 (Prospective cohorts).

**PARTICIPANTS:** Over 31,000 admissions to the intensive care units (ICUs) at two hospitals. Additionally, 777 patients with COVID-19 patients were used for prospective validation. Patients who were placed on mechanical ventilation within four hours of their admission were excluded.

**MAIN OUTCOME(S) and MEASURE(S):** Electronic health record (EHR) data were extracted on an hourly basis, and a set of 40 features were calculated and passed to an interpretable deep-learning algorithm to predict the future need for mechanical ventilation 24 hours in advance. Additionally, commonly used clinical criteria (based on heart rate, oxygen saturation, respiratory rate, FiO_2_ and pH) was used to assess future need for mechanical ventilation. Performance of the algorithms were evaluated using the area under receiver-operating characteristic curve (AUC), sensitivity, specificity and positive predictive value.

**RESULTS:** After applying exclusion criteria, the external validation cohort included 3,888 general ICU and 402 COVID-19 patients. The performance of the model (AUC) with a 24-hour prediction horizon at the validation site was 0.882 for the general ICU population and 0.918 for patients with COVID-19. In comparison, commonly used clinical criteria and the ROX score achieved AUCs in the range of 0.773 – 0.782 and 0.768 – 0.810 for the general ICU population and patients with COVID-19, respectively.

**CONCLUSIONS and RELEVANCE:** A generalizable and transparent deep-learning algorithm improves on traditional clinical criteria to predict the need for mechanical ventilation in hospitalized patients, including those with COVID-19. Such an algorithm may help clinicians with optimizing timing of tracheal intubation, better allocation of mechanical ventilation resources and staff, and improve patient care.

## INTRODUCTION

The novel coronavirus 19 (COVID-19) pandemic has strained global healthcare systems^1^ and supply of mechanical ventilation^2^, as approximately 3%–17% of hospitalized patients require invasive mechanical ventilation^3–6^. There is a major concern that the supply of mechanical ventilators is insufficient for certain regions^7,8^. Appropriate triage and identification of patients at high risk for respiratory failure may help hospital systems better guide resource allocation and cohorting of patients^8,9^. Additionally, identification of patients who may need intubation allows healthcare providers to prepare for endotracheal intubation (e.g. by moving the patient to a negative pressure room), thereby preventing an emergent procedure that is inherently high risk and aerosol-generating^10–13^. Related to fears of contamination, many providers decided to intubate early on the assumption that patients would eventually need mechanical ventilation so as to avoid ‘crash intubation’^14^. Others have called for more judicious use of mechanical ventilation, and to avoid high positive end-expiratory pressure (PEEP) in poorly recruitable lungs, which tends to result in severe hemodynamic impairment and fluid retention^15^. Both patient self-inflicted lung injury and ventilator-associated lung injury could potentially exacerbate lung inflammation and biotrauma^16^. As such, objective and consistent methods to determine who and when to intubate^17^, how to optimize treatment parameters, and when to safely extubate patients are needed to lower the long-term complications and mortality rate in this very sick patient population.

Current scoring systems that predict respiratory failure and need for mechanical ventilation are limited by small sample size and have low predictive power^18^. Frontline providers have called for urgent development of new warning systems for patients likely to fail conservative management and require mechanical ventilation^19^. Prior studies utilizing deep learning based algorithms have been shown to improve diagnostic accuracy and predict outcomes across a variety of clinical scenarios^20–25^. Such algorithms can interpret and make useful predictions from large and dynamic data available in the electronic health record (EHR). There are no reliable models to predict the need for mechanical ventilation in patients with COVID-19, therefore we sought to utilize dynamic EHR data at hourly resolution to determine if such an approach would provide value over traditional methods such as the ROX score or simple regression-based risk scores^18^. In this study, we developed and prospectively validated a deep learning algorithm that predicts the need for mechanical ventilation in hospitalized patients, and those with known or suspected COVID-19, up to 24 hours in advance of tracheal intubation.

## METHODS

Development and reporting of the prediction model presented in this study was in accordance with the checklist provided by the transparent reporting of a multivariable prediction model for individual prognosis or diagnosis (TRIPOD) consortium^26^.

### Patient population and outcome

An observational multicenter cohort consisting of all adult patients (≥18 years old) admitted to the intensive care units (ICUs) between January 2016 and January 15, 2020 at two large urban academic health centers, the University of California, San Diego Health (UCSD) and the Massachusetts General Hospital (MGH) was considered in this study. Throughout the manuscript we refer to the respective hospital systems as the *development* and the *validation* sites. Additionally, both datasets included prospectively collected validation cohorts, involving known or suspected patients with COVID-19 between February 1st and May 4th, 2020 (due to expansion of ICU care to non-traditional floors, the MGH cohort included all hospitalized patients with COVID-19 independent of explicit indication of ICU level of care). Institutional review board approval of the study was obtained at both sites with a waiver of informed consent (UCSD #191098 and MGH #2013P001024).

Data from both sites were abstracted into a clinical data repository (Epic Clarity; Epic Systems, Verona, Wisconsin) and included vital signs, laboratory values, sequential-organ failure assessment (SOFA) scores, Charlson comorbidity index scores (CCI) index, demographics, length of stay, and outcomes. Specific inputs to the model included 40 clinical variables (34 dynamic and 6 demographic variables), which were selected based on their availability in EHRs across the two hospitals considered in our study. These included vital signs measurements (heart rate, pulse oximetry, temperature, systolic blood pressure, mean arterial pressure, diastolic blood pressure, respiration rate and end tidal carbon dioxide), laboratory measurements (bicarbonate, measure of excess bicarbonate, fraction of inspired oxygen or FiO2, pH, partial pressure of carbon dioxide from arterial blood, oxygen saturation from arterial blood, aspartate transaminase, blood urea nitrogen, alkaline phosphatase, calcium, chloride, creatinine, bilirubin direct, serum glucose, lactic acid, magnesium, phosphate, potassium, total bilirubin, troponin, hematocrit, hemoglobin, partial thromboplastin time, leukocyte count, fibrinogen and platelets) and demographic variables (for more information see eTable 1 in the Supplement). Additionally, for every vital sign and laboratory variable, the slope of change since its last measurement (Δ) was included as an additional feature. All variables were organized into 1-hour non-overlapping time bins to accommodate different sampling frequencies of available data. All the variables with sampling frequencies higher than once every hour were uniformly resampled into 1-hour time bins, by taking the median values if multiple measurements were available. Variables were updated hourly when new data became available; otherwise, the old values were kept (sample-and-hold interpolation). Mean imputation was used to replace all remaining missing values (mainly at the start of each record). To assist in model training, features in the development cohort training set first underwent normality transformations and were then standardized by subtracting the mean and dividing by the standard deviation. All other datasets were normalized using the mean and standard deviation computed from the development cohort training set.

Utilization of mechanical ventilation was defined as the first occurrence of simultaneous recording of FiO2 and Positive end-expiratory pressure (PEEP). For prediction purposes, we defined our outcome of interest as continuous mechanical ventilation for at least 24 hours or mechanical ventilation followed by death. Patients who were placed on a mechanical ventilator within three hours of admission were excluded since our model makes its first prediction at hour four of ICU admission (or hospitalization in the case of MGH COVID cohort); this allows for the collection and processing of lab samples required by the algorithm to make accurate predictions.

### Model Development and Statistical Analyses

VentNet (a two layer feedforward neural network of size 40 and 25) was trained to predict the onset of mechanical ventilation 24 hours in advance, starting from hour four into admission up to the time of mechanical ventilation or end of hospitalization. VenNet was implemented in Tensorflow, version 1.12.0, machine learning frameworks for Python, version 2.7 (Python Software Foundation). The parameters of VentNet were initialized randomly and optimized on the training data from the development cohort, using the Gradient Descent algorithm with L1-L2 regularization to avoid overfitting^27^. Model interpretability was achieved by calculating the relevance score^23^ of each input feature for every predicted risk score (see eAppendix A in the Supplement).

Within the development cohort, 10-fold cross-validation (with an 80%-20% split within each fold) was used for training and testing purposes. We report median and interquartile values of the area under Receiver Operating Characteristic (AUROC or AUC) curves (and specificity at 80% sensitivity) for the held-out testing sets within the development cohort (details on precision-recall curves are presented in the Supplement). AUROCs are reported under an end-user clinical response policy in which the model would be silenced for six hours after an alarm is fired, and correct alarms that are fired up to 72 hours prior to onset of mechanical ventilation are not penalized. The best performing model at the development site was then fixed and used for evaluation on the validation cohort, and the prospectively collected cohort of COVID-19 patients. Comparison of ROC curves was performed using DeLong’s method^28^. All continuous variables are reported as medians with 25% and 75% interquartile ranges (IQRs). Binary variables are reported as percentages.

## RESULTS

### Patient Characteristics

After applying the exclusion criteria, a total of 18,528 and 3,888 ICU patients were included in the development and validation cohorts, respectively. Patient characteristics including the percentage of ventilated patients before and after application of exclusion criteria are presented in Table 1 and eTable 2 in the Supplement. Additionally, data from 26 COVID-19 patients from the development site (UCSD) and 402 patients from the validation site (MGH) were used for prospective validation (see Table 2 and eTable 3 in the Supplement).

**Table 1:** Demographic comparisons of the UCSD and MGH general ICU cohorts.

|  | UCSD (development site) |  | MGH (validation site) |  |
| --- | --- | --- | --- | --- |
| Demographics | Non-Ventilated | Ventilated | Non-ventilated | Ventilated |
| Patients, *n* (%) | 17,723 (95.6%) | 805 (4.4%) | 3,602 (92.6%) | 286 (7.4%) |
| Age, yrs (S.D) | 61.3 [48.3 72.6] | 61.2 [48.6 71.2] | 62 [51 72] | 64 [53 74] |
| Male, *n* | 10,421 | 521 | 1,948 | 173 |
| Race, *n* |  |  |  |  |
| Caucasian | 9,659 | 440 | 2,925 | 229 |
| Black | 1,330 | 60 | 191 | 19 |
| Asian | 1,081 | 43 | 119 | 8 |
| ICU LOS, hrs (IQR) | 48.3 [26.7 95.9] | 221.5 [113.8 386.9] | 50.9 [27.2 98.0] | 183.7 [92.2 309.9] |
| CCI, # (IQR) | 3 [2 7] | 3 [1 6] | 4 [2 6] | 4 [2 6] |
| SOFA, *n* (IQR) | 0.6 [0 1.8] | 3.3 [1.9 5.1] | 0.9 [0.3 2.1] | 4.1 [2.5 6.3] |
| Inpatient mortality, *n* | 869 | 329 | 223 | 109 |
| Time from ICU admission to start of ventilation, hrs (IQR) | N/A | 20 [7.8 45] | N/A | 13 [6 33] |

S.D=standard deviation; yrs=years; LOS=length of stay; ICU=intensive care unit; IQR=interquartile range; CCI=Charlson comorbidity index; SOFA=sequential organ failure assessment
Patients were excluded if 1) their length of stay was less than 4 hours or greater than 20 days, 2) no Heart Rate was recorded during their entire stay, or 3) the start of mechanical ventilation was prior to hour four of ICU admission.

**Table 2:** Demographic comparisons of the prospective validation cohorts consisting of COVID-19 patients at UCSD and MGH.

|  | UCSD COVID-19 |  | MGH COVID-19 |  |
| --- | --- | --- | --- | --- |
| Demographics | Non-ventilated | Ventilated | Non-ventilated | Ventilated |
| Patients, *n* (%) | 16 (61.5%) | 10 (38.5%) | 343 (85.3%) | 59 (14.7%) |
| Age, yrs (S.D) | 57.6 [45.2 81.6] | 52.8 [42.3 65.9] | 65 [47 78] | 61.5 [50 73] |
| Male, *n* | 9 | 7 | 176 | 40 |
| Race, *n* |  |  |  |  |
| Caucasian | 7 | <5 | 207 | 30 |
| Black | <5 | 0 | 46 | 10 |
| Asian | <5 | <5 | 13 | <5 |
| ICU LOS, hrs (IQR) | 51.4 [37.7 128.4] | 368.7 [247.0 430.0] | 131 [87.5 230] | 258.5 [141 396] |
| CCI, # (IQR) | 4 [2.8 5.3] | 2 [1 4.3] | 3 [1 6] | 3 [1 5] |
| SOFA, *n* (IQR) | 1.3 [0 2.1] | 2.5 [0 5.4] | 0.1 [0 0.7] | 3.0 [1.6 4.7] |
| Inpatient mortality,<br>*n* | <5 | <5 | 24 | 14 |
| Time from ICU<br>admission to start of<br>ventilation, hrs (IQR) | N/A | 23 [10 63] | N/A | 49.5 [20.6 143] |

S.D=standard deviation; yrs=years; LOS=length of stay; ICU=intensive care unit; IQR=interquartile range; CCI=Charlson comorbidity index; SOFA=sequential organ failure assessment
Patients were excluded if 1) their length of stay was less than 4 hours or greater than 20 days, 2) no Heart Rate was recorded during their entire stay, or 3) the start of mechanical ventilation was prior to hour four of admission.

### Model Performance on General ICU Populations

The 10 fold cross-validated AUC on the held-out development cohort testing set at 24 hours was 0.886 [0.878 0.892] (median [IQR]), and the specificity when measured at the 80% sensitivity level was 0.824 [0.818 0.838]. We observed a drop in AUC when the prediction horizon increased from 6 hours to 48 hours (from 0.950 [0.948 0.952] to 0.845 [0.838 0.869], respectively) (See eFigure 1 in the Supplement for more details). Comparisons of the VentNet algorithm against the ROX score^18^ and a logistic regression model (Baseline model 1) based on commonly used clinical variables (namely, HR, O_2_Sat, Resp Rate, and pH) are shown in Figure 1. VentNet significantly outperformed the baseline models (p<0.001) on the development cohort testing set (AUC of 0.895 versus 0.738 and 0.769, respectively) (Figure 1, panel a). Performance of the VentNet on the external validation cohort (Figure. 1, panel b) was comparable (AUC of 0.882 versus 0.782 and 0.773, respectively). See Figure 1 (panels a-b) and eFigure 2 (panels a-b) in the Supplement for additional information, including precision-recall curves.

**Figure 1:**
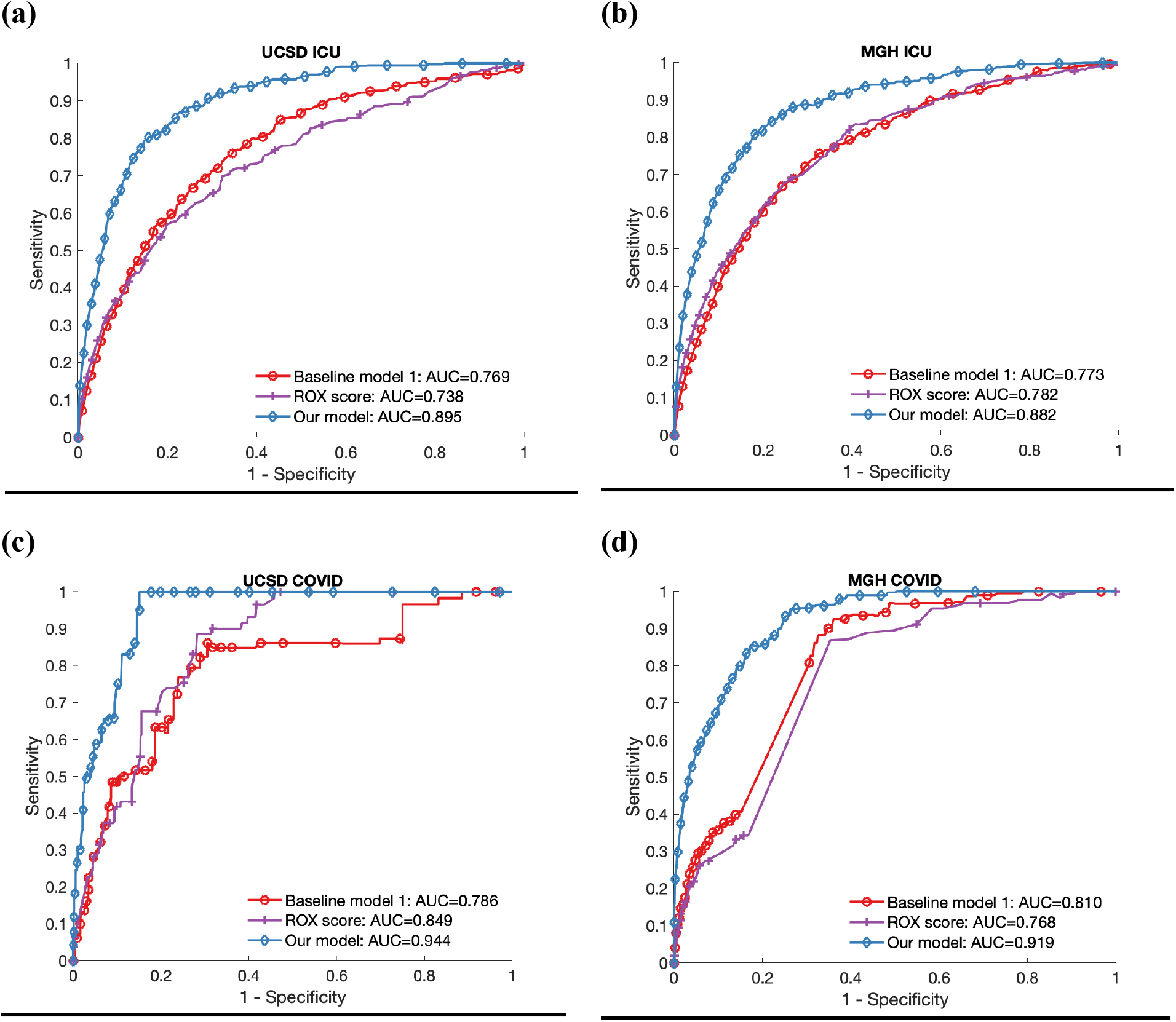
Performance of the proposed and baseline models on the development and validation ICU cohorts and the two COVID-19 prospective validation cohorts. For a prediction horizon of 24-hours, comparison of the proposed model versus two baseline models are shown on the development and validation ICU cohorts (panel a-b), and prospective validation cohorts of patients with COVID-19 (panels c-d).

Figure 2 (panels a and b) show heatmaps of the top 15 factors most commonly contributing to the increase in risk score upto 12 hours prior to intubation for the development and the validation cohorts, respectively. Some of the top predictive features included Respiratory Rate, Heart Rate, Temperature, Chloride, O_2_Sat, Platelet count, pH, and FiO2, among others. eFigure 3 in the Supplement includes an illustrative example of clinical trajectory of a patient in the ICU, as well as the respective model predictions and the top contributing factors. Note that as shown in eFigure 4 in the Supplement, a given risk factor can contribute to an increase in risk score by taking values either above or below the clinical reference range.

**Figure 2:**
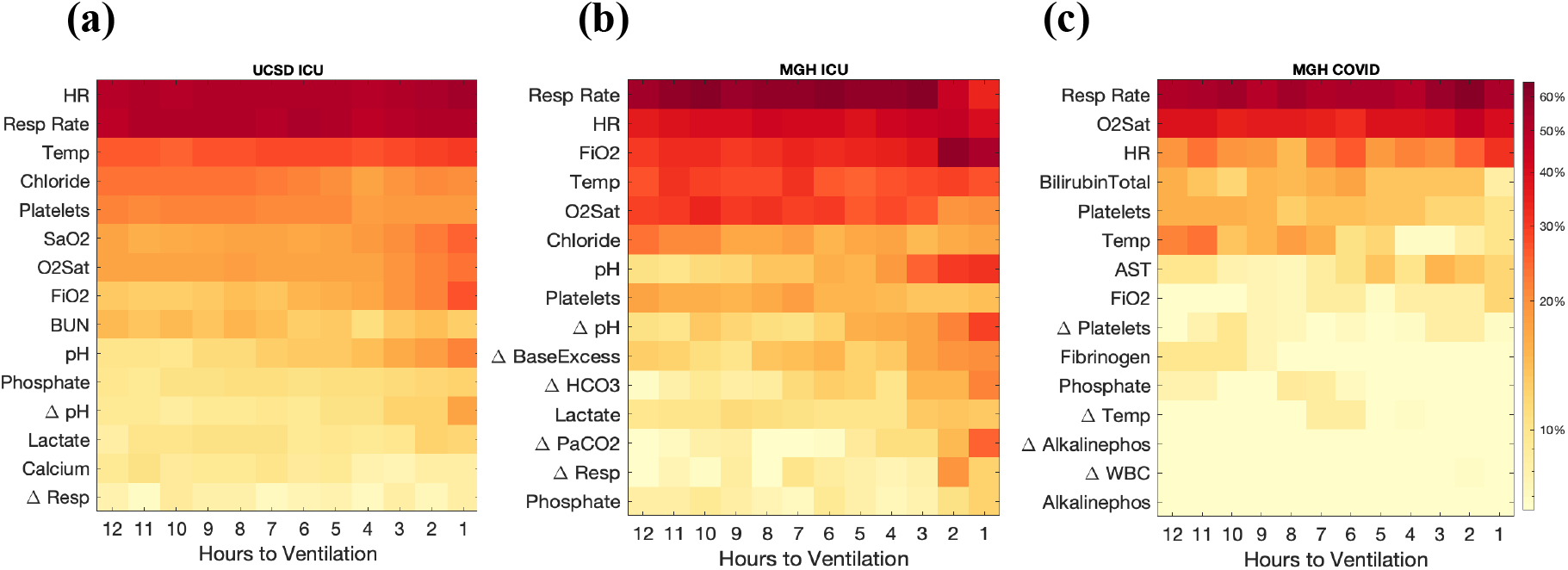
Population-level plot of top contributing factors to the increase in model risk score. The x-axis represents hours prior to onset time of mechanical ventilation. The y-axis represents the top factors (sorted by the magnitude of *relevance score*) across the patient populations at the development site (panel a), external validation site (panel b), and prospective COVID-19 cohort (panel c). Only dynamically changing variables are shown. Among the static factors, duration of time in hospital (till current time) and gender (male) were consistently among the top factors. The heat-map shows the percentage of ventilated patients for whom a given variable was an important contributor to their risk score, up to 12 hours prior to intubation. See eAppendix A in the Supplement (Interpretability section and eFigure 4 in the Supplement) for more details.

### Model Performance on COVID-19 Populations

VentNet achieved superior performance when prospectively applied to the UCSD and MGH cohorts of patients with COVID-19 (AUC of 0.943 and 0.919, respectively). The corresponding specificities measured at 80% sensitivity level were 88.8% and 84.5%, respectively. See Figure 1 (panels c-d) and eFigure 2 (panels c-d) in the Supplement for more information. Across both cohorts, performance of the VentNet was significantly higher than the ROX score and the Baseline model 1 (p<0.001; see Figure 1 and eFigure 2 in the Supplement for more details).

Figure 2 (panel c) shows a heatmap of the top 15 factors most commonly contributing to the increase in risk score upto 12 hours prior to intubation for the COVID-19 cohort at the validation site. In addition to features listed above, other factors frequently contributing to the risk score in the COVID-19 population included Total Bilirubin, Aspartate Aminotransferase (AST), Fibrinogen, and Phosphate, among others. Figure 3 includes an illustrative example of the clinical trajectory of a COVID-19 patient, as well as the respective model predictions and the top contributing factors.

**Figure 3:**
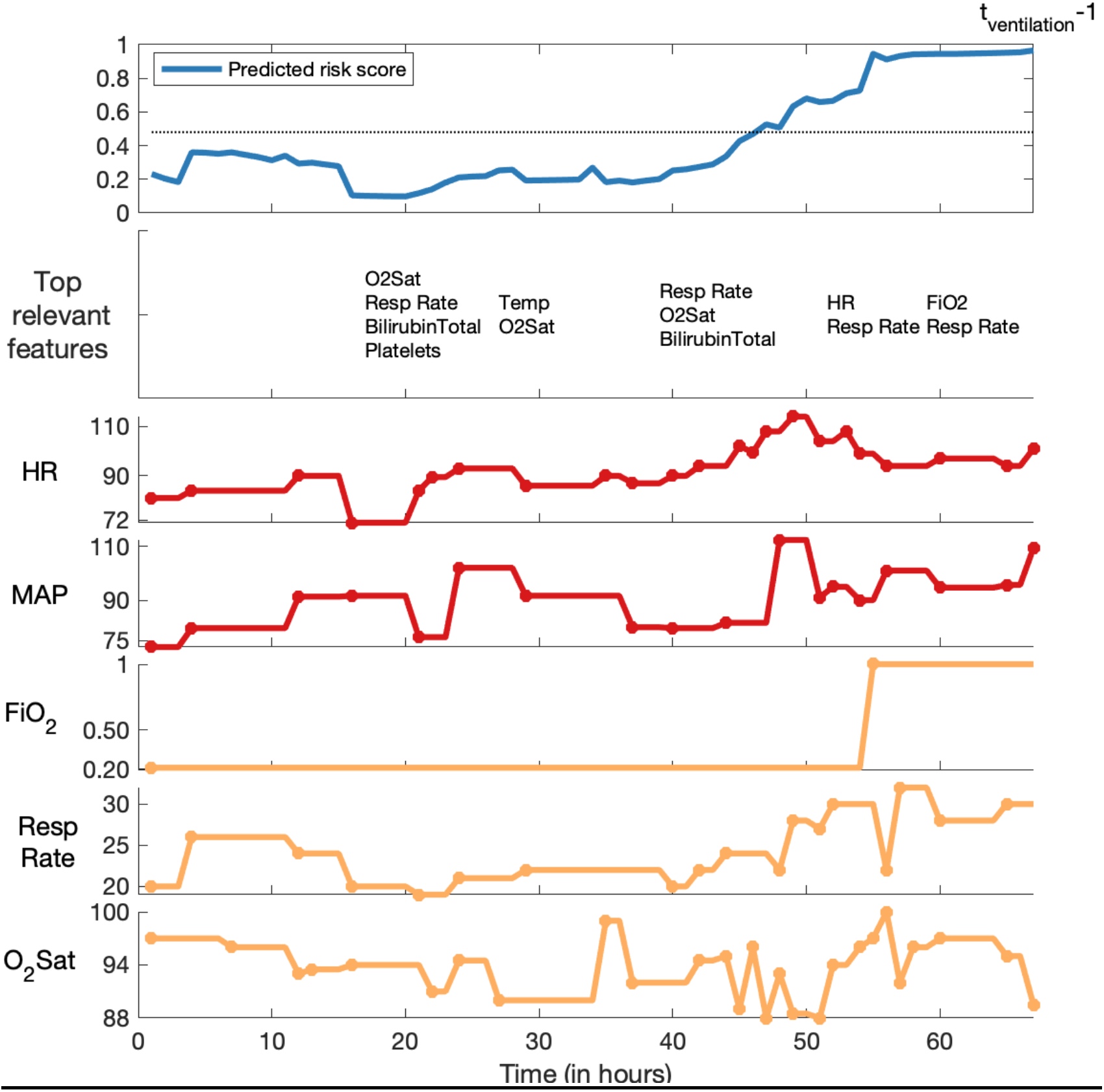
An illustrative example of a patient’s trajectory over a 67 hour window preceding intubation. The proposed algorithm crossed the prediction threshold around hour 45, roughly 24 hours prior to the onset time of mechanical ventilation. This 54-year-old female with a history of hypothyroidism presented with fevers, chills, muscle aches, fever, sore throat, cough, and anosmia. She was admitted to the hospital for hypoxemia and a chest x-ray showing basilar patchy opacities present in the emergency department. She later tested positive for COVID-19. Her oxygen requirements and work of breathing increased with a marked drop in oxygen saturation around hour 50. On the afternoon of the third day (hour 65) of hospitalization, she developed rapidly progressive respiratory failure, was intubated and diagnosed with acute respiratory distress syndrome (ARDS). For clarity, the top relevant features are shown every five hours under the estimated risk scores.

## DISCUSSION

We demonstrated that a high-performing deep learning model (AUC0.88) can predict future need for mechanical ventilation 24 hours in advance using commonly accessible EHR data. We externally validated all findings in patients from a separate academic center, as well as in two prospective cohorts of patients with COVID-19 (See Figure 1). Since the proposed model can inform healthcare providers of the most relevant features contributing to the need for mechanical ventilation (see Figures 2 and 3), it provides an interpretable algorithm to aid clinicians with optimizing timing of tracheal intubation, better allocation of resources, and improving patient care.

The COVID-19 pandemic has placed important strains on the healthcare system as the surge and long tail of critically ill patients continues to impact resource availability^1^. Despite having the highest number of ventilators and critical care beds per capita among developed countries, mechanical ventilation in the United States is still a finite resource^7,8^. Frontline providers in the pandemic noted that traditional risk stratification tools such as MEWS and quick sequential organ failure assessment (qSOFA) score are inadequate to accurately predict respiratory failure in patients with COVID-19^29^. Additionally, physicians have attempted to predict respiratory failure with simple scoring systems, yet such models have not been validated in patients with COVID19 (e.g. ROX index). To our knowledge, this is the first study to demonstrate robust performance of a deep learning algorithm for early prediction of the need for mechanical ventilation in patients hospitalized with COVID-19.

Our findings are important for a number of reasons. First, we have developed and externally validated an interpretable deep learning algorithm that predicts the need for mechanical ventilation using commonly accessible clinical variables. Such findings could be used to facilitate optimal triage, more timely management, and resource utilization. Second, we have shown with high predictive value the ability of our algorithm to function in different geographic settings in the United States and in varying cohorts. Third, our model used a sequential predictive approach such that ongoing clinical status was assessed to make important clinical predictions (see Figure 3 and eFigure 3 in the Supplement for illustrative examples). This strategy has advantages over a baseline assessment (e.g. MEWS and qSOFA) given the dynamic nature of critically ill patients. This approach paves the way for future implementation in real-time at the point of care. Fourth, as shown in eTable 4 in the Supplement, VentNet’s predictions do not heavily rely on a single or a handful of clinical variables and as such are more robust to data missingness. Thus, our model has both generalizability and portability and may have an impact not only on the current COVID-19 epidemic, but also in the expected “second wave” and beyond^30^.

For a 24 hour ahead prediction horizon, specificity of the model (on the MGH COVID-19 cohort) at 50% sensitivity was 96.5% (with a PPV of 35.3%) versus 98.9% (with a PPV of 39.2%) for 6 hours. In terms of model optimization one could argue the value in maximizing sensitivity, specificity or both. In particular, during the COVID-19 pandemic it has been argued that the avoidance of emergent procedures is a priority, since there is clearly a risk of viral transmission to providers and delays in intubation increases the risk of cardiovascular collapse^31,32^. Thus, a highly sensitive model may help to minimize the chance of a ‘crash’ intubation^33^ which leads to poor clinical outcomes and may put providers at risk of unnecessary viral exposure. On the other hand, a highly specific model may be used to avoid unnecessary intubation^14^, and the associated risks of ventilator induced lung injury, ventilator associated pneumonia^34^, and sedation and associated delirium^35^. Additionally, a shorter prediction horizon (e.g., 6 hour) may provide more clinically actionable information versus a longer prediction horizon (e.g., 24–72 hour) may inform population-level resource allocation.

Despite its many strengths, this study includes a number of limitations. First, we defined the need for mechanical ventilation in our EHR database based on the presence of PEEP and FiO_2_ measurements. We believe that this definition is robust based on considerable experience, but acknowledge that some mis-labeling could occur in any EHR based criteria. Nonetheless, we view such misclassification as random and do not expect any potential misclassifications would artificially improve our model’s performance. Second, more generally the proposed algorithm makes use of EHR data that was not originally designed for the analysis performed in our study. However, the superior performance of our algorithm, even in the presence of missing data, confirms its utility in a real-world clinical setting. Third, the COVID-19 pandemic has led to many changes in usual care including potentially earlier intubation, avoidance of high flow nasal cannula, and avoidance of non-invasive ventilation, among others. Thus, one could argue that the need for intubation of these patients may be driven by factors unique to this epidemic. However, our model was trained and validated with historical data from major academic centers prior to COVID-19. Thus, the high observed AUCs speak to the robustness of the model, even in the face of rapid changes in practice patterns. Fourth, one could argue that the outcome of intubation and need for mechanical ventilation is somewhat subjective and could be a function of local practices or intrinsic bias inherent in such decisions. However, our ability to predict a clinically important and hard outcome (need for mechanical ventilation) 6 to 24 hours in advance suggests the value of this model. Moreover, traditional clinical parameters (heart rate, respiratory rate, pH, oxygen saturation) used to make intubation decisions performed relatively poorly compared to our deep learning algorithm (AUC of 0.769 vs 0.895 on the development site testing cohort). Despite these limitations, we view our new findings as robust and likely to lead to important advances in the care of COVID-19 patients. Furthermore, our approach may extend beyond the COVID-19 pandemic to guide optimal clinical care using advanced analytics as applied to the general ICU population e.g. to determine timing and selecting of appropriate pharmacological therapies.

## CONCLUSION

In this two-center observational study, we demonstrate that high-performance models can be constructed to predict the future need for mechanical ventilation in hospitalized patients, including those with COVID-19. By using an open-source software, our validated algorithm is readily available for prospective studies aimed at determining the clinical utility of the proposed risk model for optimizing timing of tracheal intubation, better allocation of mechanical ventilation resources and staff, and improving patient care.

## Data Availability

Access to the computer code used in this research is available upon request to the corresponding author.

## Conflicts of interest and sources of funding

Dr. Nemati is funded by the National Institutes of Health (#K01ES025445), Biomedical Advanced Research and Development Authority (#HHSO100201900015C), and the Gordon and Betty Moore Foundation (#GBMF9052). Dr. Malhotra is a PI on NIH RO1 HL085188, K24 HL132105, T32 HL134632 and co-investigator on R21 HL121794, RO1 HL 119201, RO1 HL081823. ResMed, Inc. provided a philanthropic donation to UC San Diego in support of a sleep center. Dr. Malhotra received funding for medical education from Merck and Livanova. Dr. Mukerji is funded by the National Institutes of Health (#K23MH115812) and the Harvard Medical School Elenor and Miles Shore Foundation. Dr. Westover is supported by the Glenn Foundation for Medical Research and the American Federation for Aging Research through a Breakthroughs in Gerontology Grant; the American Academy of Sleep Medicine through an AASM Foundation Strategic Research Award; the Department of Defense through a subcontract from Moberg ICU Solutions, Inc, and by grants from the NIH (1R01NS102190, 1R01NS102574, 1R01NS107291, 1RF1AG064312). Dr. Wardi is supported by the National Foundation of Emergency Medicine and funding from the Gordon and Betty Moore Foundation (#GBMF9052). He has received speaker’s fees from Thermo-Fisher and consulting fees from General Electric. Other co-authors have declared no conflicts of interest and sources of funding.

## Supplementary Online Content

**eTable 1:** List of input variables used by the model.

**eTable 2:** Demographic comparisons of the UCSD and MGH general ICU cohorts (Overall cohorts without exclusion criteria)

**eTable 3:** Demographic comparisons of the UCSD and MGH COVID-19 cohorts. (Overall cohorts without exclusion criteria)

**eFigure 1:** The 10 fold cross validation performance of proposed model on development cohort held-out testing set at varying prediction horizons (6, 12, 24, 36, 48 hours).

**eFigure 2:** Precision-Recall Curves.

**eFigure 3:** An illustrative example of a patient’s trajectory over a 64 hour window preceding intubation.

**eAppendix A:** Interpretability

**eFigure 4:** Directionality with respect to influence of top factors contributing to an increase in the risk score (companion to Figure 2 in the main manuscript).

**eTable 4:** Summary of drop in AUC of a given model when a feature is treated as missing during evaluation.

**eFigure 1:**
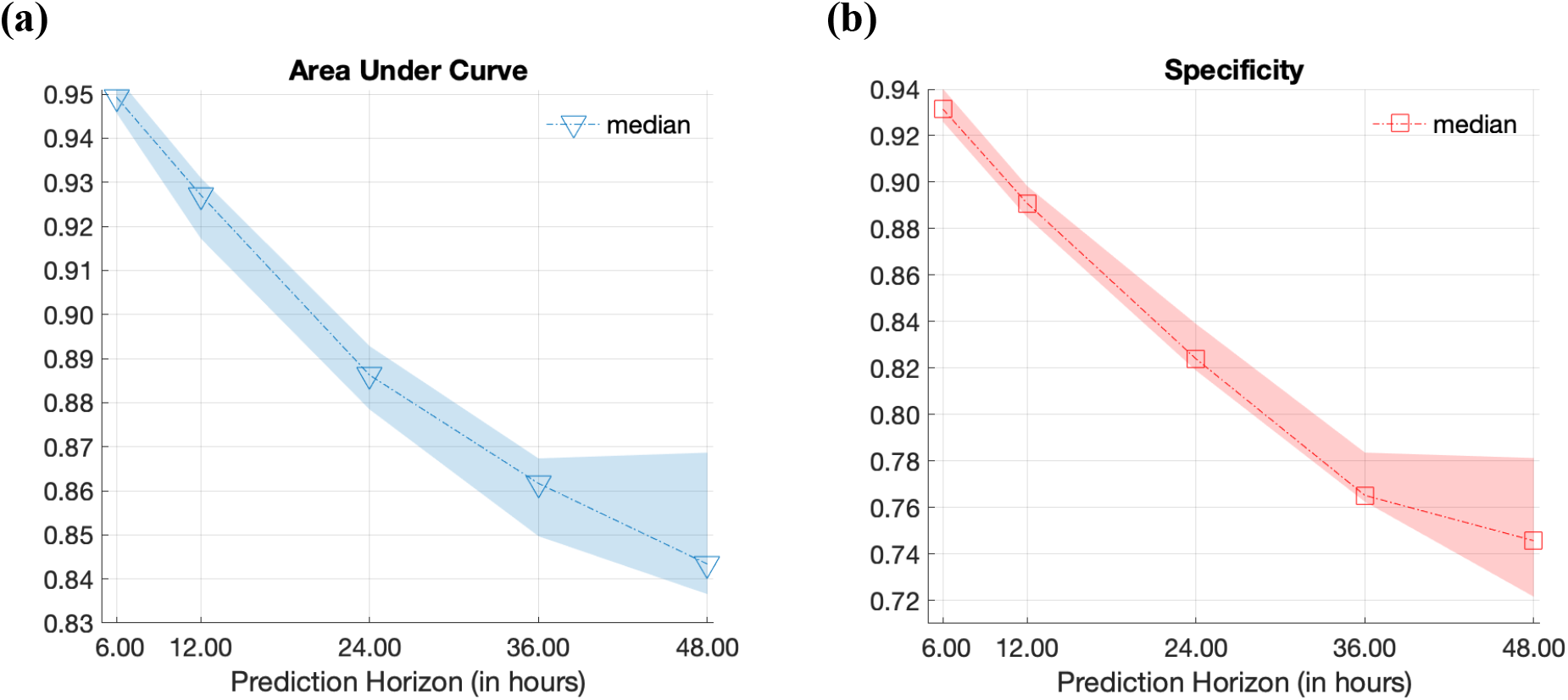
The 10 fold cross validation performance of proposed model on development cohort held-out testing set at varying prediction horizons (6, 12, 24, 36, 48 hours). Medians and Interquartile ranges (shaded area) of AUCroc and specificity (at 80% sensitivity) are shown in panels (a)- (b) as a function of prediction horizons on the held-out set of the development cohort.

**eFigure 2:**
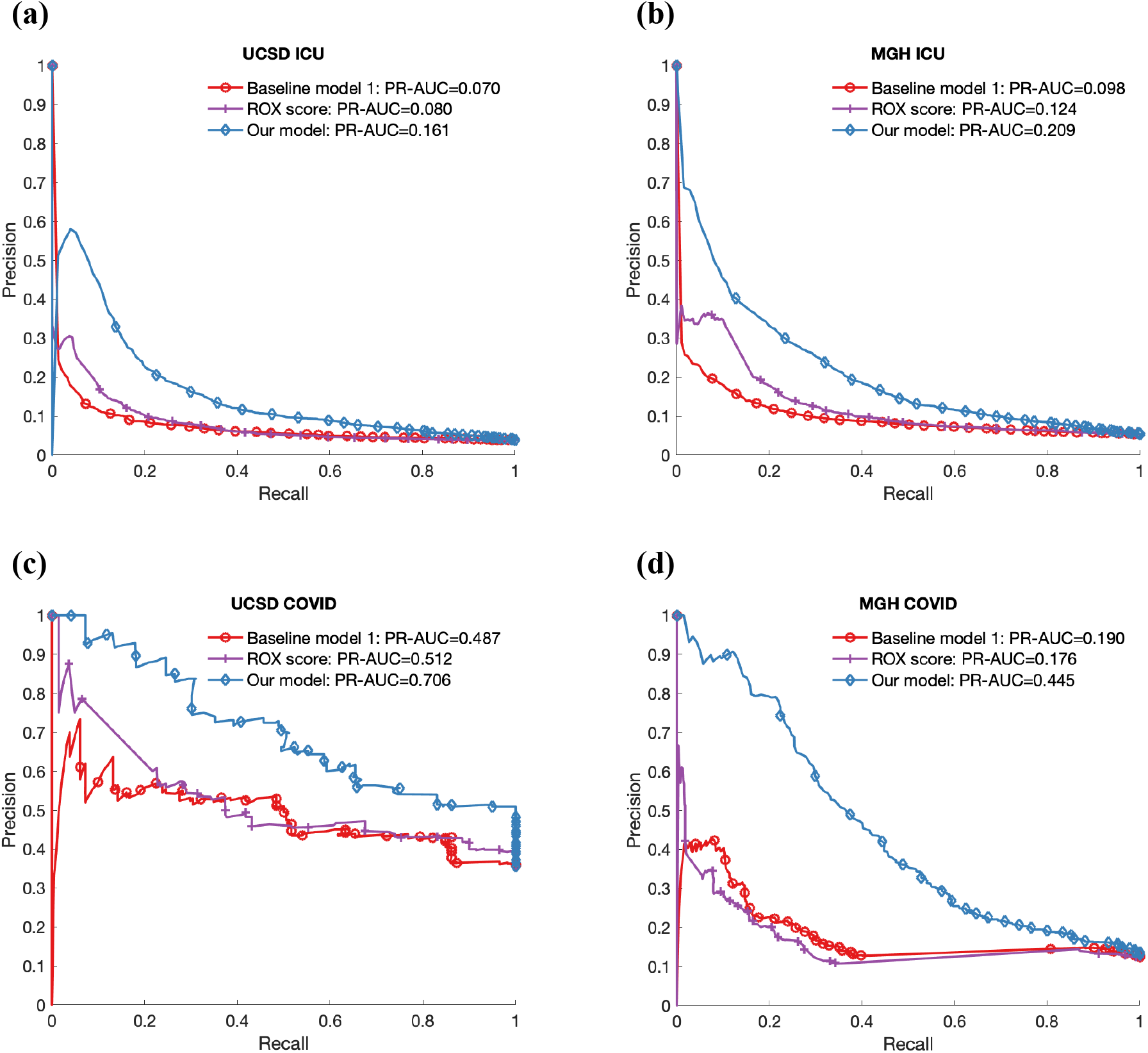
Precision-Recall Curves. Comparison of the proposed model versus two baseline models are shown on the development and validation ICU cohorts (panel a-b), and prospective validation cohorts of patients with COVID-19 (panels c-d).

**eFigure 3:**
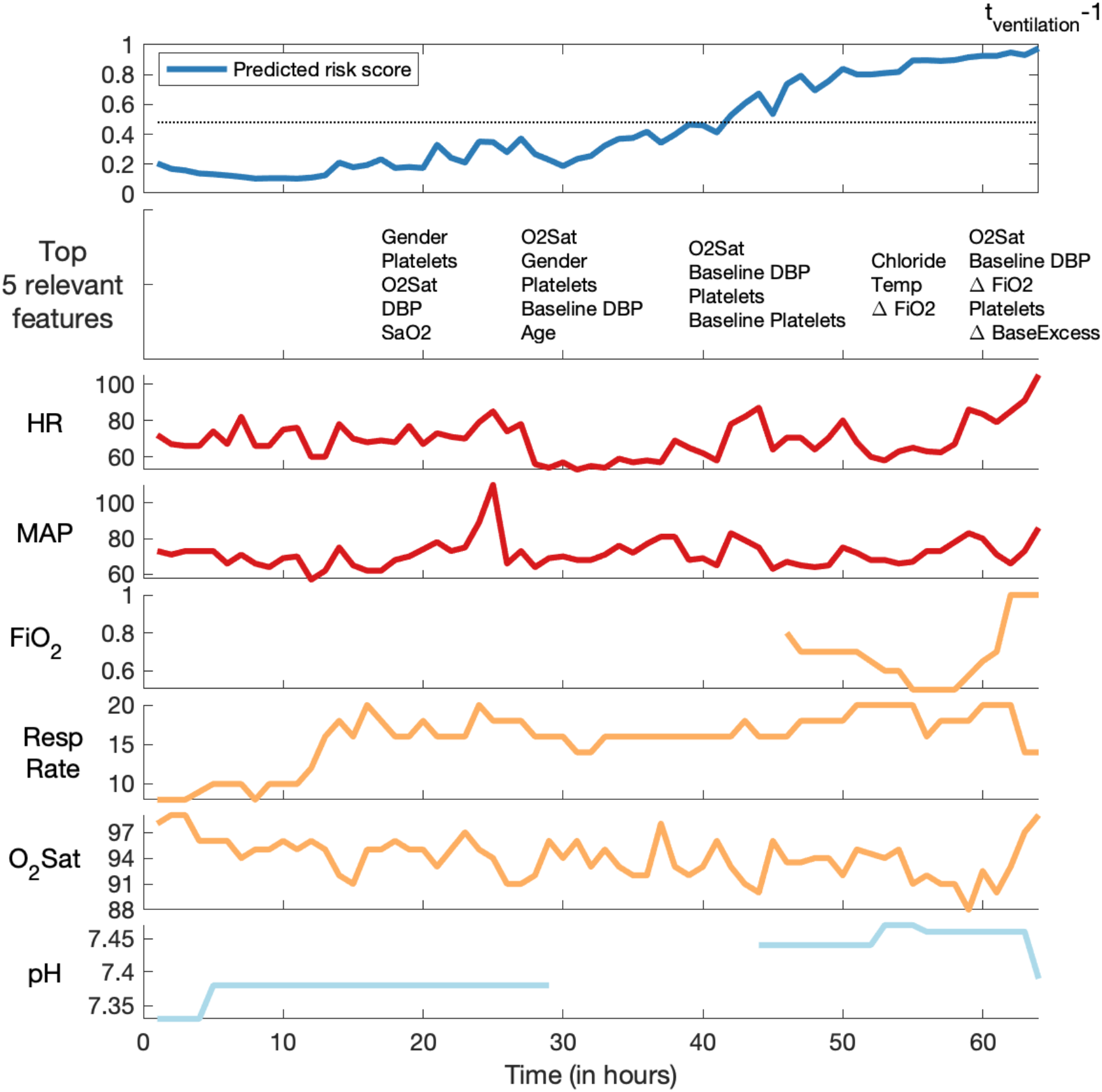
An illustrative example of a patient’s trajectory over a 64 hour window preceding intubation. This patient was a 73 year old man who developed respiratory distress, and a chest x-ray demonstrated findings concerning for aspiration vs pneumonia. He was initially treated with high flow oxygen, but ultimately required intubation and mechanical ventilation. The proposed algorithm crossed the prediction threshold around hour 40, roughly 24 hours prior to the onset time of mechanical ventilation. Notably, at hour 45 the patient was placed on 80% supplementary oxygen. Attempts to reduce the amount of supplementary oxygen within the proceeding hours resulted in a sharp drop in O_2_Sat to 88%. For clarity, the top relevant features are shown every five hours under the estimated risk scores.

**eFigure 4:**
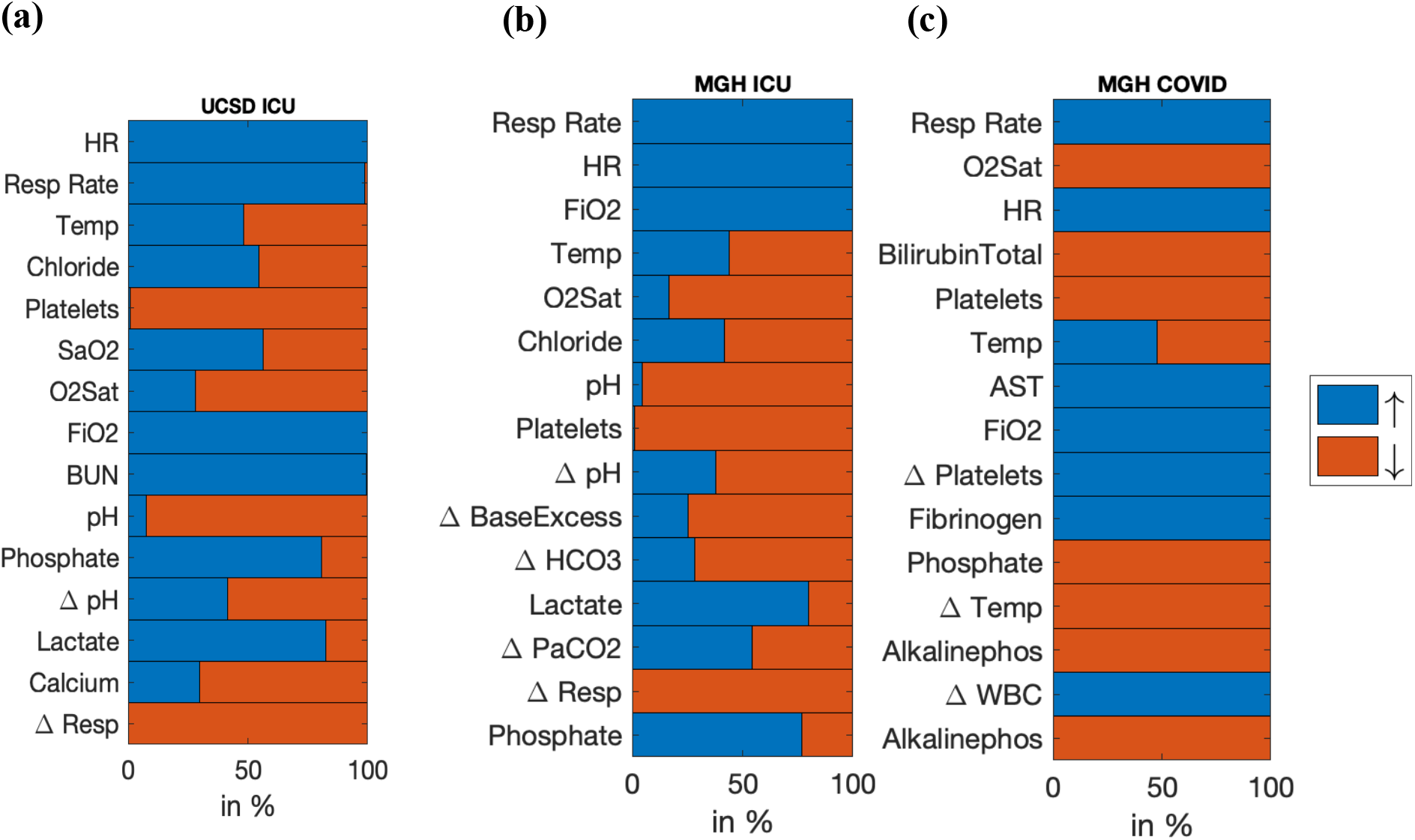
Directionality with respect to influence of top factors contributing to an increase in the risk score (companion to Figure 2 in the main manuscript). A key advantage of nonlinear models is their ability to model U-shaped risk profiles. For instance, out of all instances that temperature contributed to an increase in risk for ventilation within our various cohorts, roughly 50–60% was due to abnormally high values of temperature (color-coded as blue) and 40–50% was due to an abnormally low value of temperature (color-coded as red). Traditionally used linear models (such as logistic regression) cannot adequately capture such risk profiles. Note, these findings need to be interpreted in the context of multiplicative interactions among the risk factors, such as age and immune system deficiency. Notably, our cohort of patients with COVID-19 appear to be less heterogeneous in their risk profiles than the general ICU populations at our development and validation sites.

**eTable 1:**
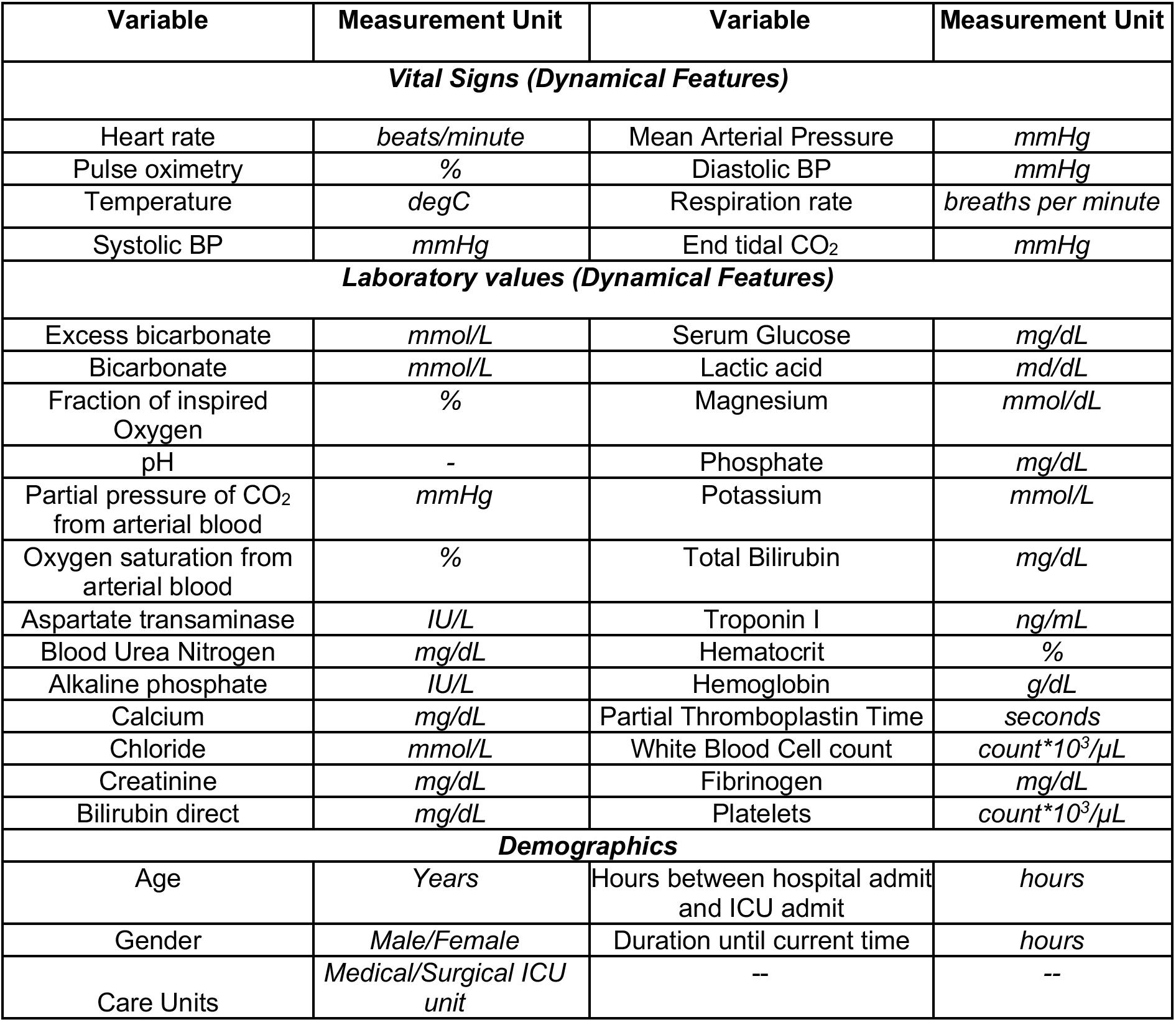
List of input variables used by the model.

**eTable 2:**
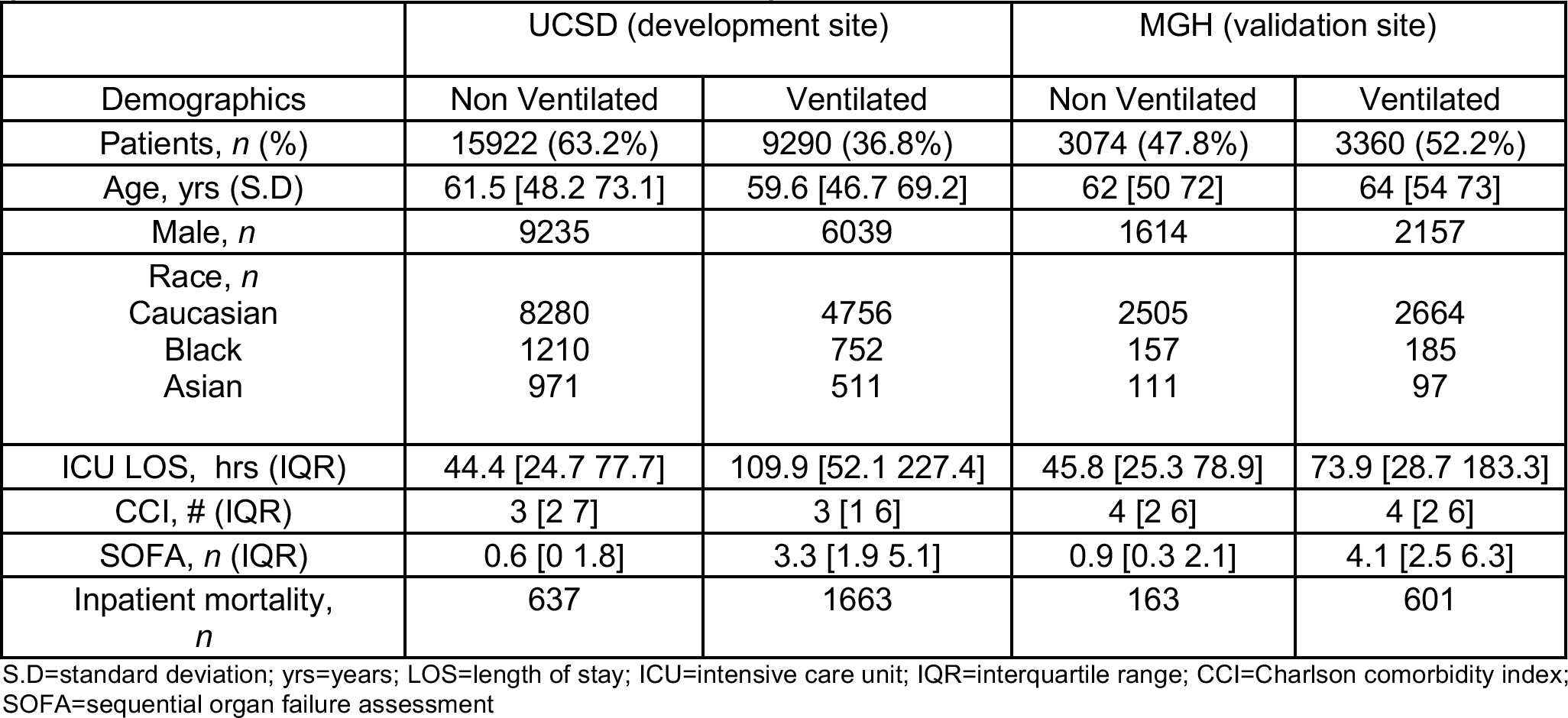
Demographic comparisons of the UCSD and MGH general ICU cohorts (Overall cohorts without exclusion criteria)

**eTable 3:**
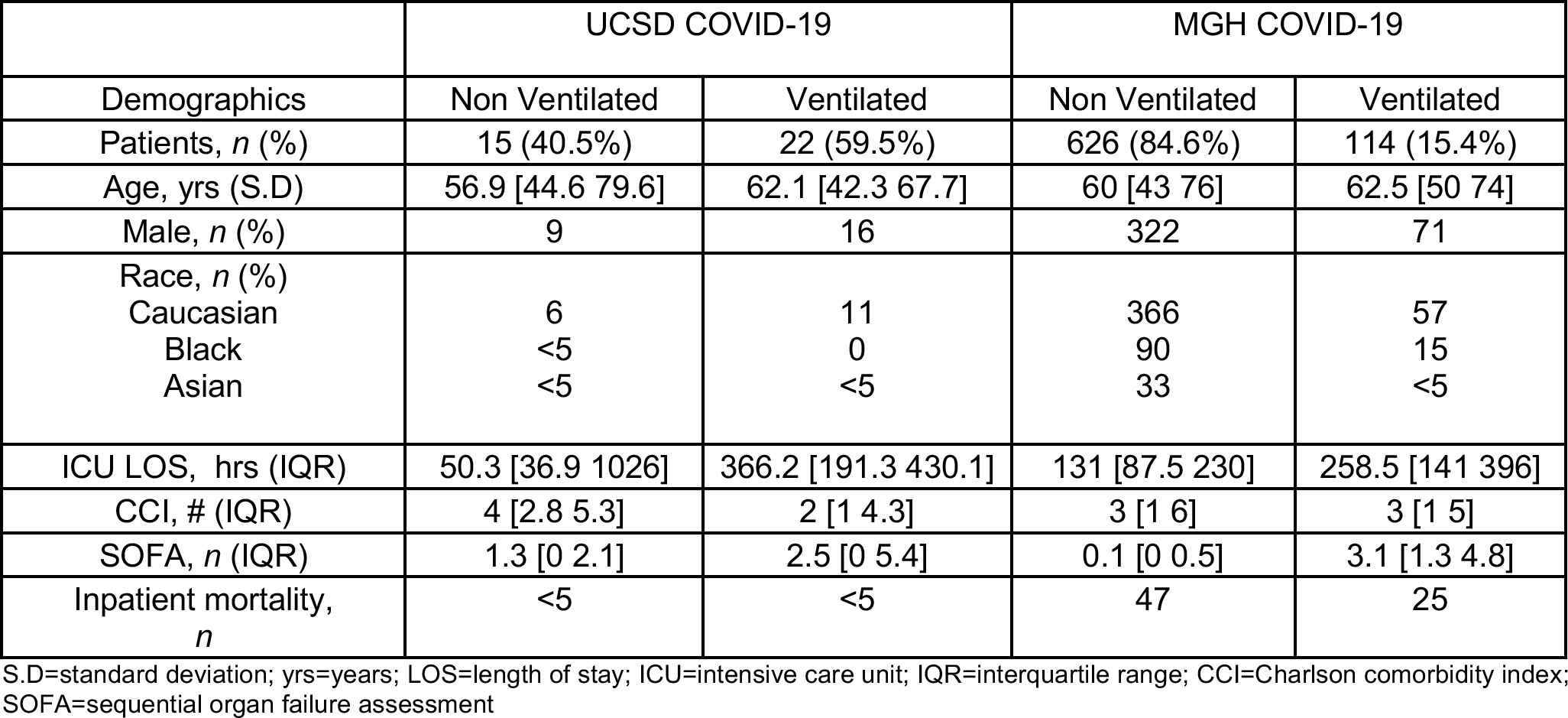
Demographic comparisons of the UCSD and MGH COVID19 cohorts. (Overall cohorts without exclusion criteria)

**eTable 4:**
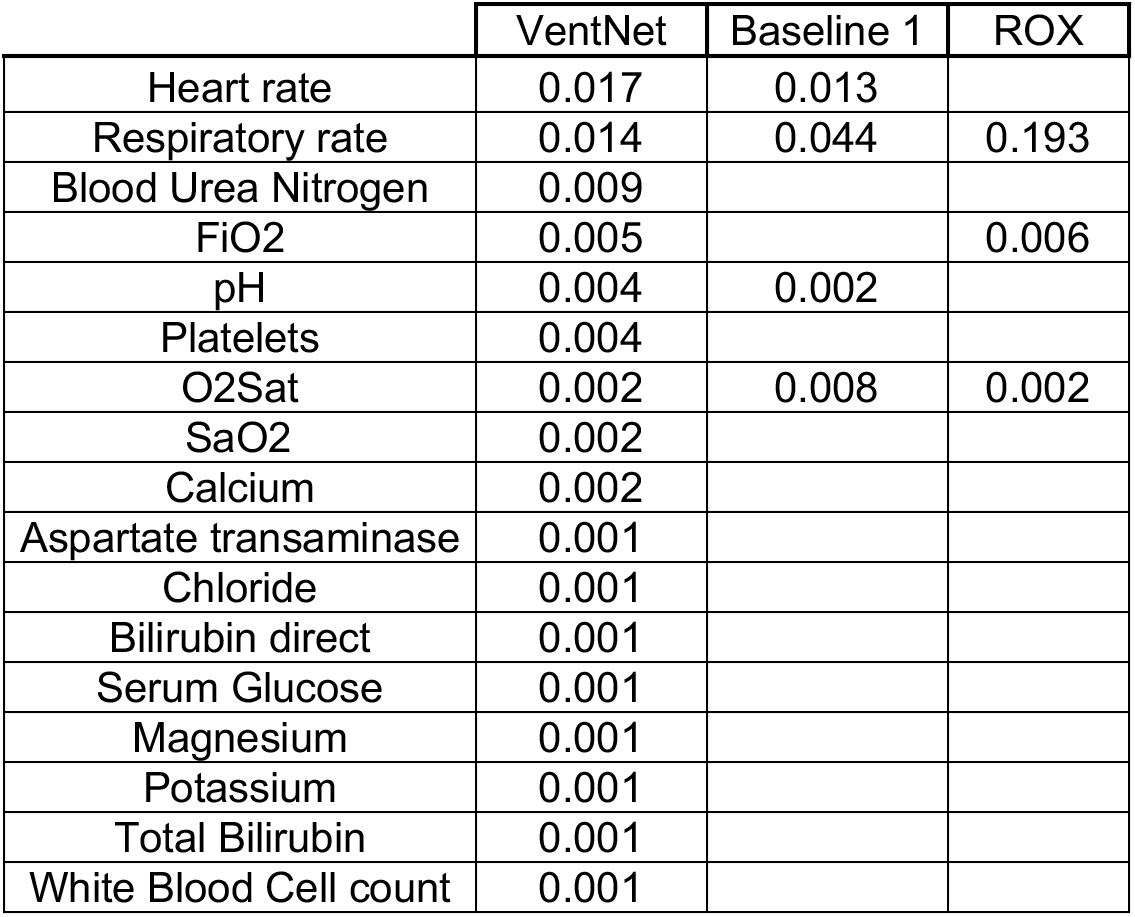
Summary of drop in AUC of a given model when a feature is treated as missing during evaluation. Results are shown for the performance of the models in the development cohort. ROX score appears to overly rely on the respiratory rate, while VentNet relies on a wider array of features to make predictions, and as such is more robust to data missingness.

## eAppendix A

### Interpretability

VentNet is uniquely interpretable wherein apart from computing the risk score, the model identifies the most relevant features contributing to the risk score as well. The importance of each feature’s contribution to the risk score is measured through a metric called *relevance score*.

To compute the relevance score, we simply take the derivative (or gradient) of the risk score with respect to all input features and multiply it by the input features. The relevance score simply says that an input feature is relevant if it is both present in the data and if the model reacts to it (the derivative term). Additionally, the direction of influence of a variable on the increase in risk score can be deduced from the sign of the input gradients (see Figure 4). In this analysis, we only extract the top contributing features with a positive relevance score.

## Notes

### Author Declarations

Institutional review board approval of the study was obtained at both sites with a waiver of informed consent (UCSD #191098 and MGH #2013P001024).

